# Exposure-Response to High PM_2.5_ Levels for Cardiovascular Events in High-risk Older Adults in Taiwan

**DOI:** 10.1101/2024.05.08.24306967

**Authors:** Shu-Ping Huang, Chien-Chou Su, Chuan-Yao Lin, Rachel Nethery, Kevin Josey, Benjamin Bates, David Robinson, Poonam Gandhi, Melanie Rua, Ashwaghosha Parthasarathi, Soko Setoguchi, Yea-Huei Kao Yang

## Abstract

**Background:** Multiple studies from countries with relatively lower PM_2.5_ level demonstrated that acute and chronic exposure even at lower than recommended level, e.g., 9 μg/m^3^ in the US increased the risk of cardiovascular (CV) events. However, limited studies using individual level data exist from countries with a wider range of PM levels to illustrate shape of the exposure-response curve throughout the range including > 20 μg/m^3^ PM_2·5_ concentrations. Taiwan with its policies reduced PM_2.5_ over time provide opportunities to illustrate the dose response curves and how reductions of PM_2.5_ over time correlated with CV events incidence in a nationwide sample.

**Methods:** Using data from the 2009-2019 Taiwan National Health Insurance Database linked to nationwide PM2.5 data. We examined the shape and magnitude of the exposure-response curve between seasonal average PM_2·5_ level and CV events-related hospitalizations among older adults at high-risk for CV events. We used history-adjusted marginal structural models including potential confounding by individual demographic factors, baseline comorbidities, and health service measures. To quantify the risk below and above 20 μg/m^3^ we conducted stratified Cox regression. We also plotted PM_2.5_ and CV events from 2009-2019 as well as average temperature as a comparison.

**Findings:** Using the PM_2.5_ concentration <15 μg/m^3^ (Taiwan regulatory standard) as a reference, the seasonal average PM_2.5_ concentration (15-23.5μg/m^3^ and > 23.5 μg/m^3^) were associated with hazard ration of 1.13 (95%CI 1.09-1.18) and 1.19 (95%CI 1.14-1.24), 1.07 (95%CI 1.03-1.11) and 1.14 (95%CI 1.10-1.18), 1.22 (95%CI 1.08-1.38) and 1.31 (95%CI 1.16-1.48), 1.04 (95%CI 0.98-1.10) and 1.10 (95%CI 1.04-1.16) respectively for HF, IS/TIA,PE/DVT and MI/ACS. A nonlinear relationship between PM_2·5_ and CV events outcomes was observed at PM_2·5_ levels above 20 μg/m^3^.

**Interpretation:** A nonlinear exposure-response relationship between PM2·5 concentration and the incidence of cardiovascular events exists when PM2.5 is higher than the levels recommended by WHO Air Quality Guidelines. Further lowering PM2·5 levels beyond current regulatory standards may effectively reduce the incidence of cardiovascular events, particularly HF and DVT, and can lead to tangible health benefits in high-risk elderly population.

## Introduction

In recent years, the global cardiology communities have increasingly recognized air pollution as a critical factors for cardiovascular health. Particulate matter with a diameter of less than 2.5 micrometers (PM_2.5_) is of particular concern due to its ability to penetrate deep into the lung tissue and enter the bloodstream, thereby posing significant risks to cardiovascular health. A growing body of evidence from multiple countries, predominantly those with relatively lower levels of PM_2.5_, has highlighted the adverse health effects associated with both acute and chronic exposure to PM_2.5_ concentrations^1–10^, even at levels below the current standards recommended by various health organizations, such as the 9 μg/m^3^ threshold set by the United States Environmental Protection Agency.

Despite the extensive research documenting the association between PM_2.5_ exposure and an increased risk of cardiovascular events^11–14^, there remains a gap in our understanding the exposure-response relationship across a wider range of PM_2.5_ concentrations in high-risk older adults. Most existing studies rely on aggregate data, which do not allow focusing on high risk groups defined by individual level factors such as having a chronic disease or age.

Taiwan presents a unique context for studying the effects of PM_2.5_ on cardiovascular events. Over recent years, Taiwan has implemented a series of policies aimed at reducing air pollution, thereby offering an invaluable opportunity to investigate the dose-response relationship between PM_2.5_ exposure and CV events in a setting where environmental policies have tangibly altered exposure levels over time. This study aims to leverage individual-level data from a nationwide sample of high-risk older adults in Taiwan to explore how seasonal exposure to moderate versus unhealthy levels of PM_2.5_ concentrations correlates with the incidence of CV events. Our study seeks to fill a critical gap in the literature by providing empirical evidence from a region characterized by a wide range of PM_2.5_ levels^15^, thereby offering a comprehensive understanding of the exposure-response curve throughout this spectrum.

The impacts of elevated levels of fine particulate matter (PM_2·5_, particles with diameter ≤ 2·5 µm) on health outcomes have been continuously and extensively studied. In recent years, studies examining the relationship between PM_2·5_ and health outcomes have utilized individual-level healthcare databases, deepening our understanding of the association between PM_2·5_ exposure and mortality as well as various hospitalizations including cardiovascular (CV) events.

## Methods

### Data Sources

Our study used data from the Taiwan National Health Insurance Database (NHID) covering the period from 2009 to 2019. The NHID is a comprehensive health insurance program that provides coverage for 99·9% of the country’s population. The database includes beneficiary registries, ambulatory care claims, inpatient claims, and prescription dispensing claims from pharmacies. Diagnosis codes based on the International Classification of Diseases, Ninth Edition, Clinical Modification (ICD-9-CM) and Tenth Edition, Clinical Modification (ICD-10-CM) were used to identify outcomes and comorbidities, along with National Health Insurance Codes identifying procedures and drug treatment. A more detailed description of the NHID has been reported elsewhere.^16^

### Cohort Definition

The study cohort consisted of patients with chronic conditions who are at high risk for CV events, including patients with atrial fibrillation (AF), heart failure (HF), carotid stenosis, coronary artery disease (CAD), cancer, peripheral vascular disease (PVD), cerebrovascular accidents (CVA), and venous thromboembolisms (VTE), as well as those who underwent recent total hip or knee arthroplasty (THA or TKA). These conditions were identified from inpatient records based on at least one code within the 12 months preceding the index date. The index date referred to the first outpatient or inpatient medical record between 2009 and 2018 in the NHID for individuals aged 65 years or older. For THA/TKA, the index date was determined as the first hospital discharge date by a post-orthopedic or major surgery inpatient code (81·54, 81·51). A detailed definition of the high-risk cohort is available in Table S-1.

### Cardiovascular Events

Cardiovascular (CV) events were defined as events with at least one inpatient diagnosis identified by an ICD-9/10 code in the first or second position. The five CV events measured in the study were acute coronary syndrome (including myocardial infarction) (ACS/MI), ischemic stroke/transient ischemic attack (IS/TIA), atrial fibrillation (AF), deep vein thrombosis (DVT), and heart failure (HF). Specific criteria and details of the CV events definitions are provided in Table S-2 of the Supplemental Material. The date of the outcome event was recorded as the first date of diagnosis after the index date. Censoring occurred due to death, end of the study period, or loss of National Health Insurance eligibility.

### Seasonal PM_2·5_ Exposure

Seasonal average PM_2·5_ concentrations were obtained from the Data Bank for Atmospheric and Hydrologic Research and linked to the NHID based on postcode. The seasons of Winter, Spring, Summer, and Fall were defined as occurring from December to February, March to May, June to August, and September to November, respectively. Nationwide estimates of seasonal average PM_2·5_ concentrations were calculated based on hourly data collected at 85 monitoring stations. Hourly PM_2·5_ concentrations were then averaged within each combination of postcode and seasons to obtain postcode-level seasonal average concentrations. For areas with no monitoring station, the PM_2·5_ concentration at the nearest monitoring station was used.

To identify each patient’s postcode of residence, we employed a residence estimation algorithm developed by Lin et al.,^17^ which had been validated against census data (Pearson correlation coefficient 0·960). The rule of residence estimation was based on the classification of beneficiaries in the NHI program and the location of hospital or clinic visits. This estimation was composed of four steps: (1) if the employment status of a beneficiary belongs to a certification requiring a professional license (such as accountant, lawyer, health care personnel), farmers, fishermen, veterans, or low-income citizens, whose residence can be designated as identical to the one registered in the NHI registry, as professional practice or veteran, and social benefit is restricted to the location of a city or county. The employment status can be identified from the database of beneficiary registry with the code numbers of “2 (62 prior to 2006),” “3,” “5,” or “6”; (2) for those who did not belong to the above categories, we retrieved the ambulatory care for upper respiratory tract infection (URI) in NHID, and assigned the location of hospital/clinic they visited as their residence area; (3) if the persons had no records for ambulatory URI, we checked for other ambulatory visits in the hospital/clinic and selected the one with highest visits as their residence location; (4) for those who had no claims record of ambulatory care other than the six defined chronic illnesses, the residence information in the most recent registry was considered to be their residence location. The residence estimation algorithm is shown in Figure S-1, which shows the validated results of residence estimation positive predictive value (PPV) by linking NHID and the National Health Interview Survey (NHIS). The overall residence estimation PPV was 84.3%.^18^

### Covariates

Demographics including age, sex, and insurance premium (a proxy for socioeconomic status) were obtained from the NHID. Information on the presence or absence of baseline comorbidities and health behaviors was obtained using outpatient and inpatient diagnosis codes from the NHID. These variables were considered as time-invariant confounders (see Table S-3 in the Supplemental Material). Measures of healthcare utilization during the baseline period including the number of hospitalizations, total hospitalization days, number of emergency department visits, number of outpatient visits, and number of medications dispensed were also taken from the NHID. Region of residence, season, and year were treated as time-varying confounders.

### Statistical Analysis

Descriptive statistics were used to summarize baseline characteristics, with continuous variables presented as means with standard deviations (SD) and categorical variables as counts and proportions. The incidence of CV events was calculated as the number of cases divided by the number of individuals in the cohort by season.

Age- and sex-adjusted incidence was estimated using Poisson regression. Marginal structural Cox models (MSM), which adjust for confounding through inverse probability of exposure weights (IPWs), were fit separately for each type of CV events hospitalization in order to estimate the effect of PM_2·5_ on the time to the first event. Following the approach used by our prior study^17^ the MSM in this study estimated the effect of PM_2·5_ exposure at a specific time point per season rather than the cumulative exposure history up to that point, as in standard MSM. Random forest models fit to the PM_2·5_ measurements, with all confounding variables as predictors, were used to estimate the IPWs for each person-season. The estimated IPWs were stabilized and truncated at the 1^st^ and 99^th^ percentiles. To adjust for death as a competing event, censoring-by-death weights (CDWs) were also estimated by fitting a random forest to an indicator of death, with PM_2·5_ and the confounders as predictors. CDWs were also stabilized and truncated. The final inverse probability weights deployed in the MSMs were generated by multiplying the IPWs and CDWs for each person-season.

To illustrate the exposure-response curves, we first conducted an inverse probability weighted Cox regression with a restricted cubic spline on PM_2·5_. For the restricted cubic spline models, we plot the estimated spline curves across the range of observed PM_2·5_ concentrations to evaluate the shape of the exposure-response curve for each outcome, with 15 μg/m^3^ – the Taiwan Environmental Protection Agency air quality standard concentration of PM_2·5_ – used as the reference PM_2·5_ level. To quantify the impact of PM_2.5_ on CV events at moderate vs. high levels, we estimated hazard ratio of average seasonal PM_2.5_ level of moderate (15-23.5ug/m3) and high ( > 23.5 ug/m3) using PM_2.5_ level < 15ug/m3 as reference.

All statistical analyses were conducted using SAS 9·4 (SAS Institute Inc., Cary, NC, USA) and R software (version 4·2·1). The study was approved by the National Cheng Kung University Hospital Institutional Review Board (IRB no: B-EX-108-056).

## Results

Among 373,402 older adults with high-risk conditions in Taiwan, the average age was 73·6 years, and 57·7% of cohort members were female with the majority with hypertension (72%) or osteoarthritis (58%). The mean PM_2·5_ concentration observed in our study was 28·3 μg/m^3^ (standard deviation 17·4 μg/m^3^) (Table 1). The median PM_2·5_ concentration during the study period was 23·5 μg/m^3^ with minimum of 0.1 μg/m^3^ and maximum of 72.65 μg/m^3^ with significant seasonal variation (Figure 1).

**Figure 1.**
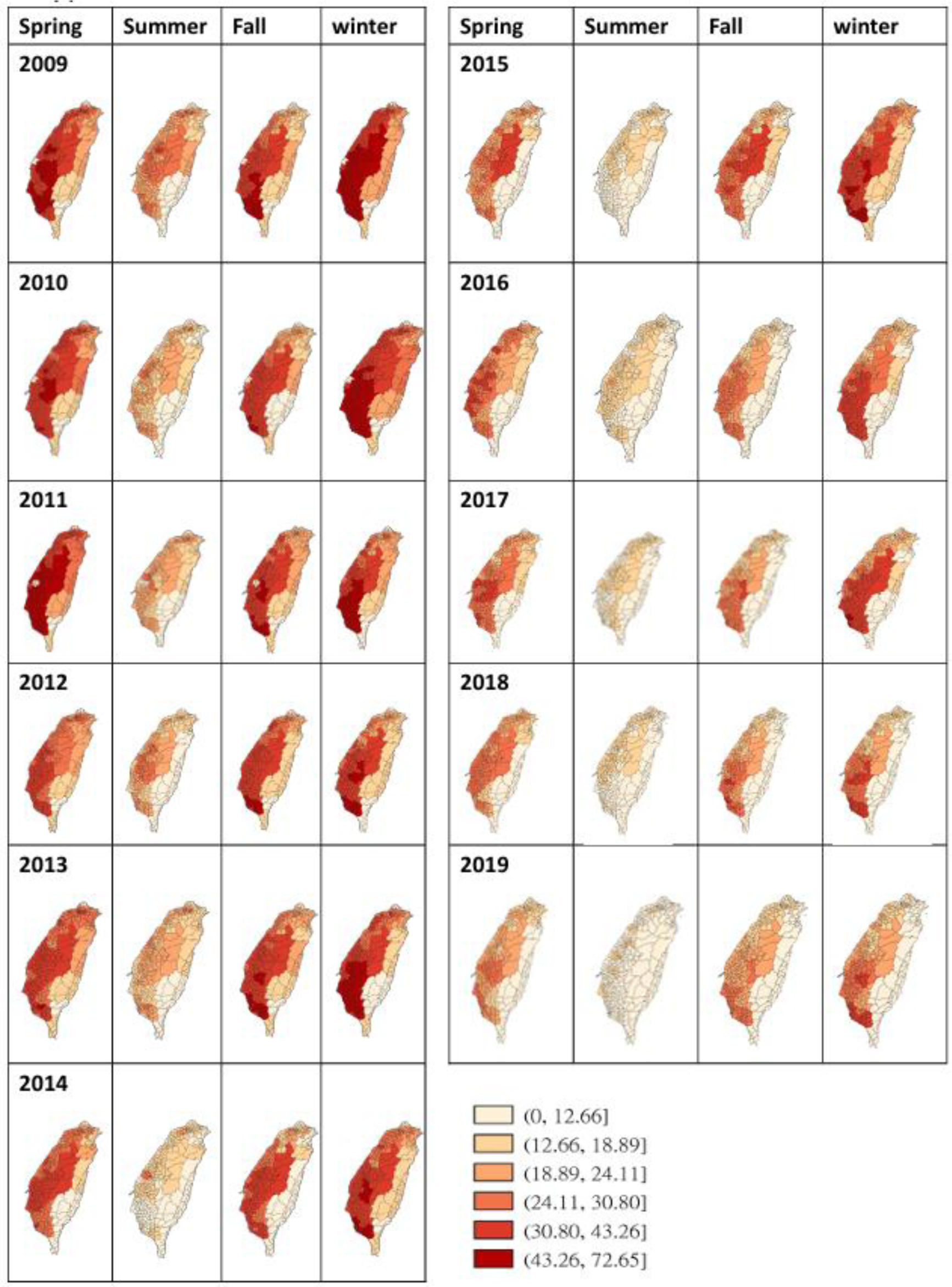
Maps of postcode-level seasonal average PM_2.5_ concentrations (ug/m3) over the study period.

**Table 1.** Baseline characteristics of the high CV events risk cohort (N=373,402).

|  | Mean (or N) | SD (or %) |
| --- | --- | --- |
| **Exposure** |  |  |
| PM<sub>2.5</sub> µg/m<sup>3</sup> | 28.31 | 17.41 |
| **Individual Characteristics** |  |  |
| Age | 73.61 | 7.22 |
| Male | 157948 | 42.3 |
| **Area-Level Characteristics** |  |  |
| Population density | 2494.95 | 3187.88 |
| National Health Insurance premium | 22907.41 | 20382.02 |
| **Individual Comorbidities, Behavioral Risk Factors, and Health Service** |  |  |

| **Measures** |  |  |
| --- | --- | --- |
| Smoking | 838 | 0·22 |
| Obesity | 1345 | 0·36 |
| Diabetic nephropathy | 23400 | 6·27 |
| Diabetic neuropathy | 13813 | 3·7 |
| Diabetes retinopathy | 7321 | 1·96 |
| Diabetes with peripheral circulatory disorders | 5328 | 1·43 |
| Diabetes overall (uncomplicated + complicated; Type I or II) | 129975 | 34·81 |
| Hypertension | 272363 | 72·94 |
| Hypercholesterolemia | 116000 | 31·07 |
| Stroke | 68782 | 18·42 |
| Transient ischemic attack | 20681 | 5·54 |
| Myocardial infarction | 13096 | 3·51 |
| Heart failure | 70066 | 18·76 |
| Chronic kidney disease | 30167 | 8·08 |
| Renal insufficiency (non-CKD related) | 22847 | 6·12 |
| Stable angina | 29725 | 7·96 |
| Unstable angina | 18426 | 4·93 |
| Other ischemic heart disease | 93347 | 25 |
| Peripheral vascular disease | 10502 | 2·81 |
| Atrial fibrillation | 33885 | 9·07 |
| Other dysrhythmias | 42987 | 11·51 |
| Cardiac conduction disorder | 3844 | 1·03 |
| Other cardiovascular diseases | 23647 | 6·33 |
| Asthma | 34232 | 9·17 |
| Chronic obstructive pulmonary disease | 38641 | 10·35 |
| Chronic pulmonary disease (includes<br>bronchiectasis) | 59130 | 15·84 |
| Obstructive sleep apnea | 194 | 0·05 |
| Pneumonia | 47709 | 12·78 |
| Acute renal disease | 9222 | 2·47 |
| Chronic renal insufficiency | 43336 | 11·61 |
| Edema | 29899 | 8·01 |
| Liver disease | 65157 | 17·45 |
| Hypothyroidism | 5560 | 1·49 |
| Hyperthyroidism | 4487 | 1·2 |
| Depression | 22211 | 5·95 |
| Anxiety | 43833 | 11·74 |
| Sleep disorder | 69053 | 18·49 |
| Dementia | 28312 | 7·58 |
| Psychosis | 4353 | 1·17 |
| Obesity | 1640 | 0·44 |
| Smoking | 2178 | 0·58 |
| Alcohol use | 2763 | 0·74 |
| Drug abuse | 807 | 0·22 |
| Congenital anomalies of the heart and<br>great arteries | 1055 | 0·28 |
| Cardiomyopathy | 3957 | 1·06 |
| Ventricular arrhythmia | 1887 | 0·51 |
| Disorders of mineral and electrolyte imbalance | 32119 | 8·6 |
| Diarrhea | 23142 | 6·2 |
| Hyperparathyroidism | 731 | 0·2 |
| Angioneurotic edema | 184 | 0·05 |
| Diseases and dysfunctions of GI/GU | 183849 | 49·24 |
| Food allergy | 1265 | 0·34 |
| Drug allergy | 4025 | 1·08 |
| Glaucoma | 1635 | 0·44 |
| Myocarditis | 183 | 0·05 |
| Neutropenia | 3311 | 0·89 |
| Cancer | 102415 | 27·43 |
| Epilepsy | 2902 | 0·78 |
| HIV | 58 | 0·02 |
| Anemias | 63297 | 16·95 |
| Any mental disorder | 110321 | 29·54 |
| Bipolar disorder | 2093 | 0·56 |
| Dyskinesia | 15 | 0 |
| Diverticulosis of intestine | 2063 | 0·55 |
| Pancreatitis | 2649 | 0·71 |
| Osteoarthritis | 215556 | 57·73 |
| Disc disorder | 29296 | 7·85 |
| Hip fracture | 8411 | 2·25 |
| Self-harm | 26 | 0·01 |
| Number of outpatient visits | 36·09 | 21·82 |
| Number of hospital visits | 1·74 | 1·61 |
| Days hospitalized | 15·82 | 25·54 |
| Number of emergency department visits | 1·01 | 1·65 |
| Number of medications dispensed | 13·58 | 5·79 |

Figure 2 shows the time series of seasonal PM_2·5_ concentrations and the age- and sex-adjusted incidence rates of the five CV events (DVT, ACS/MI, HF, IS/TIA, and AF). PM2.5 levels showed clear seasonal patterns and declined gradually over time. Incidence of HF and IS/TIA were high among Taiwanese older adults with a range of 0·2-2·7% and 0·2-1%, respectively. The age and sex-adjusted incidence of HF, ACS/MI, DVT/PE and IS/TIA have also declined over time with HF and ACS/MI showing clear seasonal patterns synchronous with PM2.5 concentrations.

**Figure 2.**
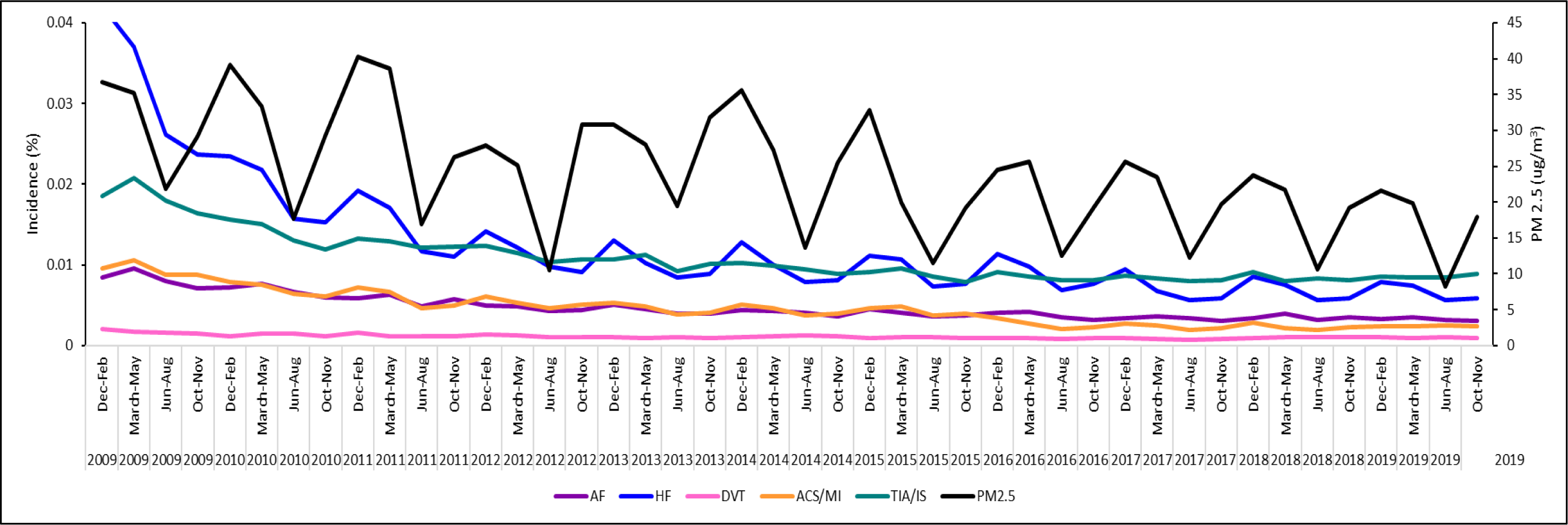
Seasonally adjusted incidence of cardiovascular events. Taiwan started to set the PM2.5 threshold regulation in 2012. It has been set at 15 µg/m3 from the beginning and remains unchanged. Taiwan began implementing the Air Pollution Prevention and Control Action Plan in December 2017 to reduce the national annual average PM2.5 concentration from 20 µg/m3 in 2016 to 12.4 µg/m3 in 2022. (For a detailed report, please refer to the press release of the Ministry of Environment https://www.moenv.gov.tw/en/AD00D74F752160/b7a33398-f7e6-4938-b868-36204b1c8270).

Restricted cubic spline curves showed that the risk of HF, DVT, IS/TIA, and ACS/MI all increased as PM_2·5_ concentrations increased (Figure 3). For DVT and HF, the slope of the curve was steeper at concentrations below 20 μg/m^3^ and leveled off at higher concentrations. The curve for IS/TIA was linear up to at approximately 45 μg/m^3^ before it started to level off. For ACS/MI, the slope was less steep at lower concentrations but more steep at higher PM_2.5_ concetration, approximately at 35 μg/m^3^. We observed no between PM_2·5_ concentrations and AF.

**Figure 3.**
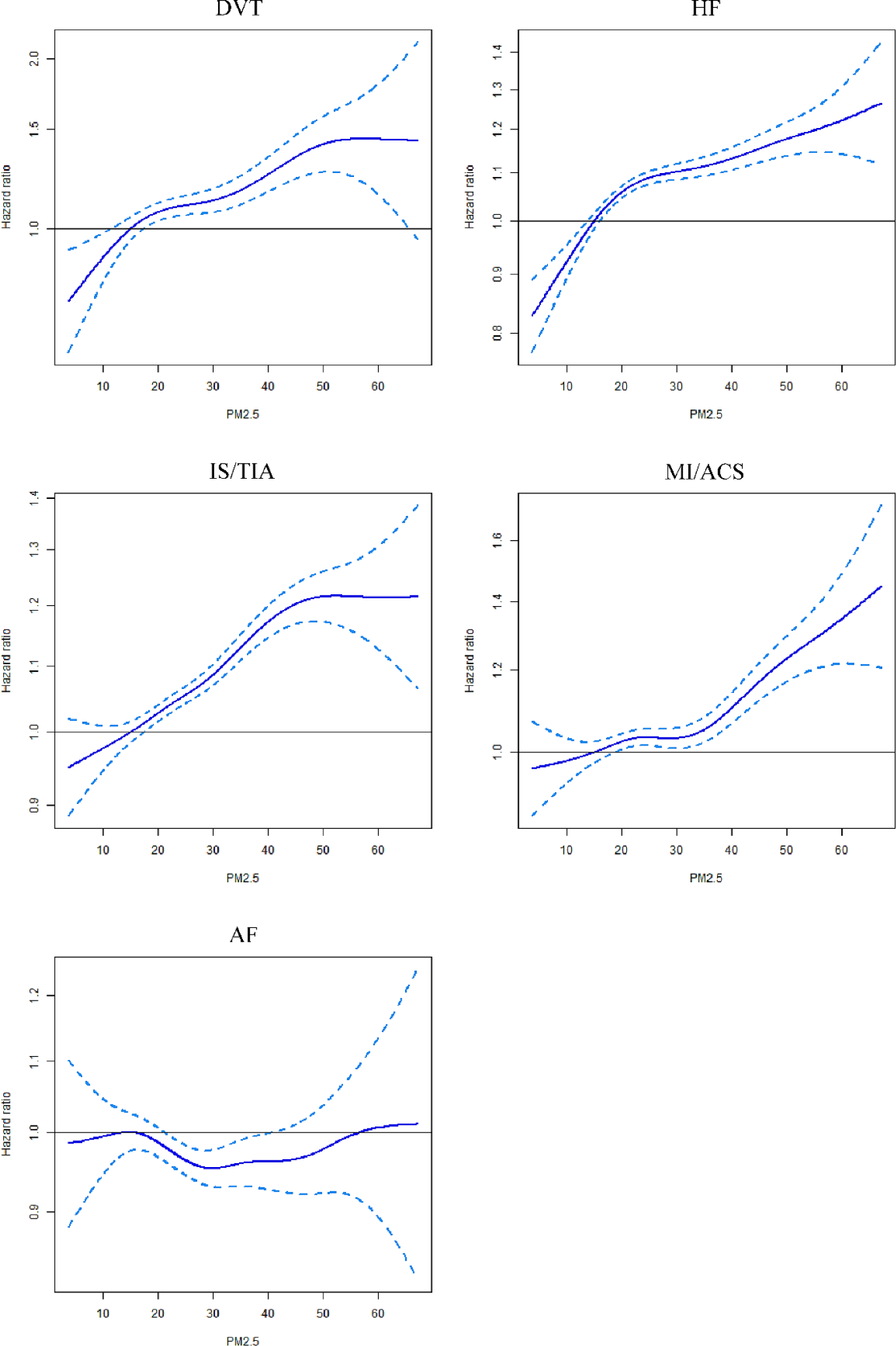
Hazard ratios estimated by MSM with restricted cubic spline Cox models to evaluate associations between PM_2·5_ and cardiovascular events (reference point 15 μg/m^3^).

Compared to the PM_2.5_ concentration <15 μg/m^3^ (the current Taiwan regulatory standard) as a reference, the adjusted hazard ratio (HR) for moderate (15-23.5μg/m^3^) on HF, IS/TIA, and PE/DVT were 1.13 (95%CI 1.09-1.18), 1.07 (95%CI 1.03-1.11), and 1.22 (95%CI 1.08-1.38), respectively (Figure 4) but was much smaller and not statistically significant for MI/ACS (HR=1.04, 95%CI 0.98-1.10). HR for PM2.5 on AF was almost null. The adjusted HR for high (> 23.5μg/m3) on HF, IS/TIA, PE/DVT and MI/ACS were 1.19 (95%CI 1.14-1.24), 1.14 (95%CI 1.10-1.18), 1.31 (95%CI 1.16-1.48) and 1.10 (95%CI 1.04-1.16), respectively. These findings were consistent with the exposure-response curves illustrated by cubic spline curves in Figure 3.

**Figure 4.**
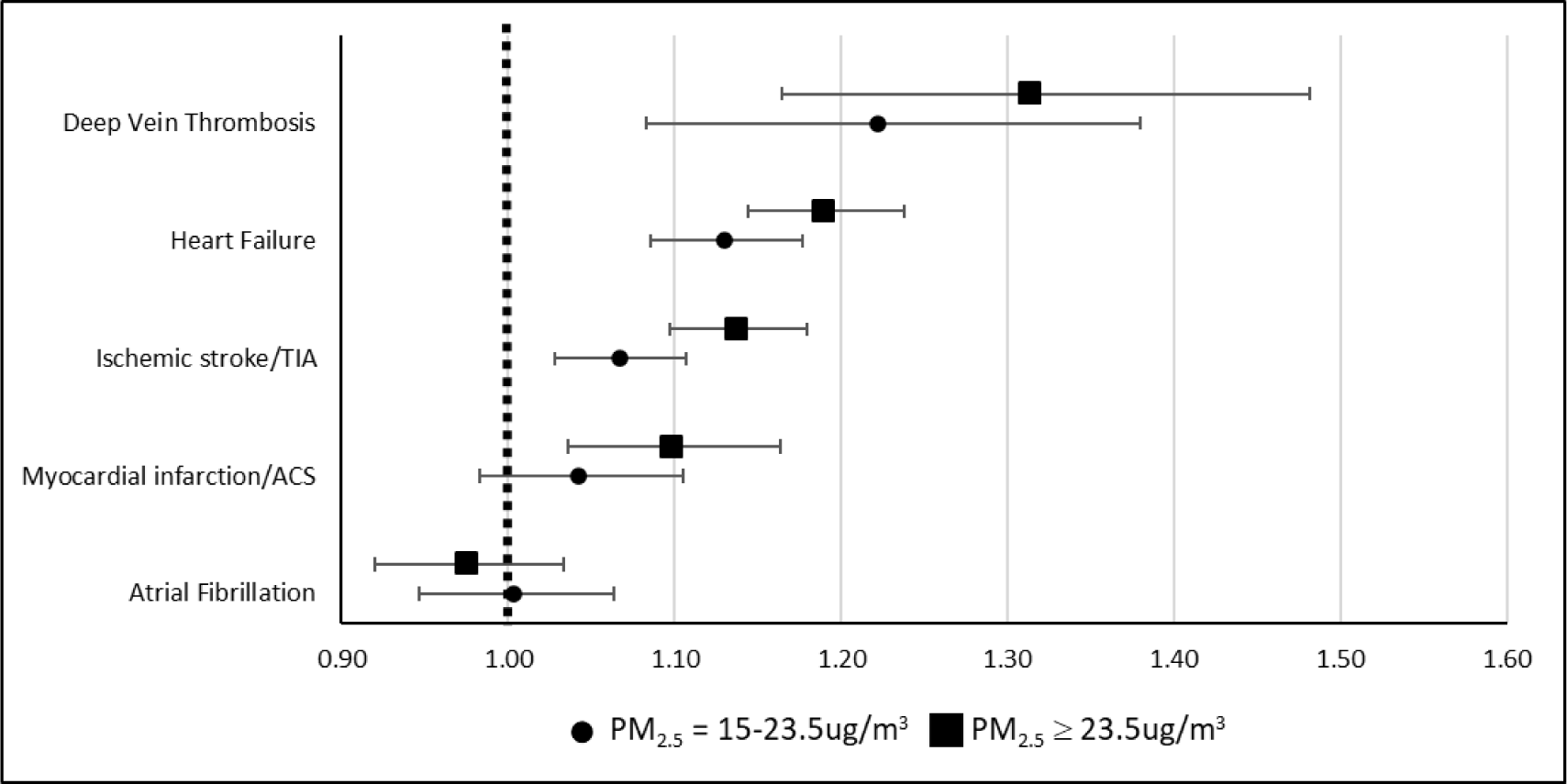
Forest plot of adjusted hazard ratio of average seasonal PM_2.5_ level (< 15ug/m3, 15-23.5ug/m3 and > 23.5 ug/m3) as categorical variable for each CV events outcome. Average seasonal PM2.5 level < 15ug/m3 was used as reference.

## Discussion

In the national sample of older adults in Taiwan, we observed a decrease over time in both air pollution (PM_2.5_) levels and the age and gender adjusted rates of CV events from 2009-2019, with HF and MI/ACS showing clear seasonal changes that matched the PM2.5 patterns. Among older adults with high-risk conditions for CV events and frequent exposure to high levels of PM_2·5_ in Taiwan, we observed non-linear exposure response relationship between PM_2·5_ and CV events. For DVT, HF, and IS/TIA, exposure to PM_2·5_ was associated with elevated risk of DVT, HF, IS/TIA with steeper slopes of the PM_2·5_ exposure-response curves at lower exposure levels. For MI/ACS, in contrast, the PM2.5 effect was more pronounced at higher exposure level.

The incidence of CV events in Taiwan and the levels of PM_2·5_ both exhibited decreasing trends over time. In previous literature, decreasing trends of incidences of HF and strokes were reported during 2010-2015^19^ and 2004-2011^20^, respectively, while AMI incidence remained constant during 2009-2015^21–22^. The patterns of our findings are consistent with results from previous studies that have reported improvements in air quality due to the implementation of various air pollution control measures in Taiwan over the past several decades.^23^ For instance, Taiwan’s Air Pollution Control Act, which has been continuously strengthened since its introduction in 1975, has played a significant role in regulating and reducing air pollution levels. The Air Pollution Prevention and Control Strategy, implemented since 2017, has further contributed to air quality improvement through a combination of incentives and constraints.

American Heart Association (AHA) scientific statement on PM air pollution and cardiovascular disease in 2010 concluded that PM_2.5_ increases the risk of cardiovascular events and death, but there are few quantitative summaries available that synthesize and compare the magnitudes of these effects. A meta-analysis which included 42 studies, focus on long-term PM_2.5_ exposure and risks of ischemic heart disease and stroke events shows that long-term PM_2.5_ exposure is associated with increased risks of IHD mortality, cerebrovascular mortality, and incident stroke. However, this study does not provide the level of PM_2.5_ and assumes that the effect of PM_2.5_ exposure on CV risk is linear. ^24^

Our spline models enabled us to characterize the shape of the relationships between PM_2·5_ and each CV events across exposure levels ranging from 0.1 μg/m^3^ to 72.65μg/m^3^. For DVT, HF, MI/ACS and AF, a plateau was observed in the curve between levels of 20 μg/m^3^ and 40 μg/m^3^. At PM_2·5_ concentrations below 20μg/m^3^, increasing PM_2·5_ exposure was associated with increased risk of DVT, HF, MI/ACS and AF. Conversely, above 20 μg/m^3^, there were smaller positive but not statistically significant associations between PM_2·5_ concentrations and most CV events. This provides important evidence about the complex non-linear relationships between PM_2·5_ exposure and CV events at higher PM_2·5_ concentrations. Our study showed biphasic concentration-response relationships between PM_2·5_ levels and CV events. This pattern is consistent with results from studies conducted in areas with high PM_2·5_ levels,^25–28^ as well as with the American Heart Association’s consensus that associations between PM_2·5_ concentration and CV events risk appear to be monotonic below 15 μg/m^3^.^11^

Our results differ somewhat from those of Lo et al., who investigated the association between PM_2·5_ and cardiopulmonary disease in the Taiwanese population.^29^ They found that each 10 μg/m^3^ increase in 5-year average exposure to PM_2·5_ was associated with a 4·8% increased risk of incident ischemic heart disease (95% confidence interval [CI] −3·3, 13·6) and a 3·9% increased risk of incident stroke (95% confidence interval [CI] −2·9, 11·1). However, they did not detect any evidence of non-linearity for most outcomes examined, with the exception of ischemic heart disease. However, Lo et al. used 5-year average PM_2·5_ concentrations whereas we used more granular, seasonal average values. PM_2·5_ concentrations vary substantially by season and by year in Taiwan as shown in our analyses. Thus, seasonal averages reflect patient exposures to fine PM more precisely than 5-year averages. Also, the study populations are different between the two studies; Taiwan National Health Interview Survey (62,694 individuals from general older adults) in Lo et al. vs. NHID (373,402 individuals who are at high risk for CV events). Our study population was larger, more nationally representative but also focused on high risk individuals.

The mechanisms by which PM_2·5_ influences cardiovascular outcomes involve both direct and indirect pathways. Animal studies have demonstrated that PM_2·5_ can be directly distributed through the bloodstream to remote target organs, leading to local oxidative stress, inflammation, and atherosclerotic plaques, ultimately resulting in thrombus formation.^30^ Indirectly, PM_2·5_ deposited in the lungs can trigger oxidative stress and inflammation, leading to increased circulating levels of pro-inflammatory cytokines, a known risk factor for atherosclerosis.^11,25^ A study conducted in Beijing found that exposure to higher levels of air pollution led to acute inflammatory and prothrombotic response in the early lag period, followed by a gradual decrease in effect over time, potentially due to compensatory mechanisms.^26^

Our study utilized seasonal average exposures in part to provide actionable insights for policy makers, clinicians and individuals. This approach was chosen in light of distinct and predictable seasonal patterns in PM_2·5_ concentrations, which offer opportunities for clinicians to advise high-risk patients regarding exposure reduction during periods of elevated ambient PM_2·5_ levels, As the high-risk individuals in our study had multiple outpatient visits per season, clinicians can prescribe personal air purifiers and frequent use, and/or recommend limiting outdoor activities during seasons with high ambient PM_2·5_.^15^

Several limitations to our study should be noted. First, exposure data were obtained from fixed monitoring stations, and patient residence was indirectly estimated from claims data, introducing the potential for exposure misclassification. To mitigate this bias, we used the most frequently visited clinic as a surrogate for patient residence. Additionally, the lack of socioeconomic data in the NHID limited our ability to account for individual-level measures taken to mitigate air pollution exposure, such as the use of air purifiers or reducing outdoor exposure during periods of poor air quality. The use of air purifiers in Taiwan was about 10% between 2009 and 2014, increasing from 11% to 23% between 2015 and 2019^31^. We were able to address this limitation to some extent by using insurance premium (which reflects income) as a proxy for health behavior.

In conclusion, our study contributes to the existing knowledge regarding PM_2.5_ and cardiovascular events by demonstrating nonlinear exposure-response relationship between PM_2·5_ concentrations and incidence of CV events underscores the importance of reducing PM_2·5_ levels even below the current regulatory standard of 15 μg/m^3^ and the potential for such a policy change to effectively decrease the incidence of DVT and HF further in high-risk individuals. Accordingly, further reduction of PM_2·5_ levels beyond Taiwan’s regulatory standards (<15 μg/m^3^) may effectively reduce the incidence of DVT and HF. We also showed that the incidence of most CV events and PM_2·5_ concentrations in Taiwan fell significantly between 2009 and 2019, supporting the potential effectiveness of comprehensive air pollution control measures.

## Data Availability

The datasets generated and analyzed for the conduct of this study are not publicly available and are restricted to authorized users of the Taiwan Health and Welfare Data Science Center .

## Acknowledgments

Funding for this study was provided by the National Institute on Aging, National Institutes of Health (R01 #1R01AG060232-01A1). Gabriel D. Shapiro provided editorial support for this manuscript and was paid by Rutgers University.

## Sources of Funding

This study was funded by the National Institutes of Health (NIA R01 #1R01AG060232-01A1).

## Disclosures

The authors declare that they have no competing interests.

## Contributors

Conceptualization (AV, SS, DR), data curation (SH, CS), formal analysis (SH, CS), funding acquisition (SS), investigation (DR, RN, SS), methodology (RN, KJ, SS, CS, CL), project administration (SS, MR), resources (YY), software (CS), supervision (DR, SS, RN, BB, YY), validation (SS, BB, YY), visualization (SH, CS), writing – original draft (SH, CS), writing – review and editing (all authors).

## Notes

### Competing Interest Statement

The authors have declared no competing interest.

### Author Declarations

The study was approved by the National Cheng Kung University Hospital Institutional Review Board (IRB no: B-EX-108-056).

